# Early Prediction of Gestational Diabetes Using Integrated Cell-free DNA Features and Omics-derived Genetic Scores

**DOI:** 10.1101/2025.09.03.25334985

**Authors:** Vinh Nguyen Dao, Nhat-Thang Tran, Ta-Son Vo, Hong-Thinh Le, Thu-Ha Thi Nguyen, Quoc-Huy Vu Nguyen, Minh-Thi Thi Ha, Tam Minh Le, Diem-Tuyet Thi Hoang, Khanh-Trang Nguyen Huynh, Nhan Viet Nguyen, Chuong Canh Nguyen, Thuong Chi Bui, Xuan Thanh Nguyen, Sa Viet Le, Vinh Dinh Tran, My-Nhi Ba Nguyen, Thong Van Nguyen, Tuyet-Anh Thi Nguyen, Ba Phuoc Hoang, Trong Van Nguyen, Thuy-Ai Thuy Nguyen, Toa Tri Nguyen, Thang Duc Duong, Cuong Huy Pham, Kim-Oanh Thi Luong, Cuong Ngoc Dao, Khanh Van Hoang, Thu-Thanh Thi Huynh, Khuong Manh Nguyen, Son-Tra Thi Tran, Hoanh Trung Tran, Son Canh Nguyen, Thuy Dinh Tran, Phương Thi Lan Nguyen, Thanh Viet Pham, Kong Chi Pham, Minh Doan Thai, My-Hang Thi Truong, Hieu Ha Pham, Thanh-Thuy Thi Do, Sang Hung Tang, Hoai-Nghia Nguyen, Minh-Duy Phan, Hoa Thi Dao, Hoa Giang

## Abstract

**Background:** Gestational diabetes mellitus (GDM) affects 15.6% of pregnancies globally, with Vietnam exhibiting one of the highest prevalences at 21%. Current diagnostic approaches at 24-28 weeks limit early intervention opportunities. We developed a multi-modal machine learning framework integrating cell-free DNA (cfDNA) structural features and genetic information for early GDM prediction at 10-12 weeks of gestation in Vietnamese women.

**Methods:** We analyzed blood samples from 1,086 pregnant women (435 GDM cases, 651 controls) collected at 9-12 weeks. Two parallel analytical pathways were employed: cfDNA profiling extracting cfDNA-specific features (fragment length, end motifs, GC content, nucleosome patterns), and whole-genome imputation generating predictions for ∼19,000 omics traits. Component scores were developed using TabPFN classifier and integrated via logistic regression into a unified master score.

**Results:** Genome-wide analysis identified five omics traits with significant GDM associations: *HSD11B1*, *NEK7*, *COMMD10*, *KLRC4*, and *OCEL1*. Component score optimization revealed distinct patterns—cfDNA scores peaked at 200 features (AUC=71.53), while genetics-based scores improved with up to 2,000 omics traits (AUC=77.21). The final master score, integrating three components (gbSC_2000_, gbSC_BH_, cfSC200), achieved AUCs of 86.82 - 87.19 across validation cohorts with 70% sensitivity and 89% specificity. Addition-deletion analysis confirmed that both cfDNA and genetic components provided essential, non-redundant contributions.

**Conclusions:** This multi-modal framework demonstrates superior performance compared to single-biomarker approaches, enabling risk stratification from very low (4% GDM prevalence) to very high risk (90% prevalence). At the cutoff 0.4, the model identifies 78% of future GDM cases at 10-12 weeks while maintaining an 18% false-positive rate, potentially enabling early interventions to prevent GDM development and associated complications.

## Introduction

Gestational diabetes mellitus (GDM) affects approximately 15.6% of pregnancies worldwide, ranking as one of the most common metabolic complications during pregnancy (H. Wang et al., 2022). This condition significantly increases the risk of adverse maternal and neonatal outcomes such as preeclampsia, operative delivery, macrosomia, and birth trauma (Buchanan et al., 2012; Wicklow & Retnakaran, 2023). The consequences of GDM extend far beyond pregnancy: more than 40% of affected women develop type 2 diabetes within ten years, and their children carry lifelong elevated diabetes risk, perpetuating an intergenerational cycle of metabolic disease (Sheiner, 2020).

Current diagnostic approaches rely on glucose tolerance testing at 24-28 weeks of gestation, limiting the window for meaningful early intervention during critical developmental periods. This diagnostic timing constraint is increasingly seen as a fundamental limitation, especially since emerging evidence suggests that GDM-associated metabolic changes can be detected much earlier in pregnancy (Zhang & Yang, 2022; Zhu et al., 2022). Furthermore, current screening approaches provide only binary classification at 24-28 weeks, missing opportunities for the more nuanced risk stratification that early prediction could enable. Effective early GDM prediction requires not only high accuracy but also clear risk stratification to guide clinical decision-making—identifying women at various risk levels who might benefit from different intensities of intervention.

Cell-free DNA (cfDNA) circulating in maternal blood offers a promising non-invasive approach for early detection of pregnancy complications. The cfDNA, which comes from both maternal and placental sources, provides molecular insights into gestational processes throughout pregnancy. Recent studies have shown that deep learning models analyzing cfDNA sequencing data—including copy number variations, fragmentation patterns, and transcription start site accessibility—can predict GDM with impressive accuracy as early as 12 weeks of gestation (Tang et al., 2024; Y. Wang et al., 2023). These methods significantly outperform traditional screening approaches and enable detection weeks before the current diagnostic standards.

However, cfDNA features may represent only one component of GDM complex etiology. Emerging evidence suggests that genetic predisposition plays a substantial role in GDM development, with genome-wide association studies identifying numerous susceptibility loci across diverse populations (Pervjakova et al., 2022). The challenge lies in translating this genetic knowledge into clinically actionable prediction models, particularly given the polygenic nature of GDM where thousands of variants or genes may contribute small individual effects. Recent advances in omics prediction models offer a promising approach to capture this genetic complexity by integrating information across approximately 19,000 molecular traits, potentially bridging the gap between genomic discovery and clinical application (Xu et al., 2022). Single-biomarker approaches may be insufficient for complex diseases like GDM, where multiple biological pathways—including insulin signaling, inflammatory responses, and placental metabolism—contribute to disease pathogenesis.

Vietnam exhibits one of the world’s highest GDM prevalences at 21.0% (*Global Prevalence of Gestational Diabetes (GDM) | IDF Atlas*, n.d.). This elevated prevalence may reflect unique genetic susceptibility patterns, environmental factors, or gene-environment interactions specific to Vietnamese populations. Population-specific prediction models are increasingly recognized as essential, given that genetic risk variants and their effect sizes can vary substantially across ancestries, potentially limiting the transferability of prediction models developed in other populations. Yet Vietnam currently lacks population-specific early detection tools that account for these unique genetic and environmental factors.

In this study, we developed a comprehensive machine learning framework that integrates multiple data modalities—including cfDNA features and genetic-derived omics trait predictions—to predict GDM at 10-12 weeks of gestation in Vietnamese women. Our two-stage approach first develops specialized component scores for each data type, then combines them into a unified master score optimized for clinical risk stratification. The framework was validated using independent datasets to ensure generalizability and clinical utility. This precision medicine approach may address a critical unmet clinical need while establishing a framework for population-specific, multi-modal GDM prediction that could potentially transform prenatal care and reduce the substantial burden of diabetes-related complications in this high-risk population.

## Methodology

### Participants

This retrospective case-control study enrolled pregnant women from multiple leading maternity hospitals and clinics across Vietnam. Eligible participants had undergone non-invasive prenatal testing (NIPT) between 9^th^ and 12^th^ week of their gestation and were subsequently assessed for GDM during routine antenatal screening at 24–28 weeks, with GDM defined according to national guidelines. Women carrying fetuses with chromosomal abnormalities were excluded to reduce potential confounding from genetic anomalies.

A total of 1,086 pregnant women met the inclusion criteria and completed the study protocol. Of these, 435 women were diagnosed with gestational diabetes mellitus (GDM) based on standard screening conducted between the 24th and 28th weeks of gestation, while 651 women with normal glucose tolerance served as the control group.

### Sequencing & Imputation

All of 1086 NIPT samples were sequenced at low coverage (0.05x – 0.8x). Sequencing was performed using the AVITI platform, with detailed procedural information available in (Tran et al., 2020). Whole-genome imputation, with the exception of chromosomes X and Y, was conducted using QUILT2 (Li et al., 2024). The fetal fraction — a required input for QUILT2 — was estimated by the SeqFF program (Kim et al., 2015). Reference genotype maps were generated from the KHV genome and aligned to the hg38 assembly.

### Score construction & alignment

To construct a comprehensive GDM risk score, we first developed multiple component scores, each capturing distinct types of information available for pregnant women. These component scores were derived independently using the TabPFN classifier, a state-of-the-art AutoML algorithm for small datasets (Hollmann et al., 2023, 2025). Subsequently, these component scores were aligned onto a common risk scale through simple logistic regression, yielding a unified master score. To support this two-stage framework, the dataset was partitioned into three mutually exclusive subsets: TRAIN (n = 652; 40.34% GDM), VALIDATION (n = 217; 36.41% GDM), and DISCOVERY (n = 217; 42.86% GDM). The TRAIN set was used to develop component scores, the VALIDATION set was employed to calibrate and align them on a unified risk scale, and the DISCOVERY set was reserved for independent performance evaluation of the master score.

#### Cf-DNA score (cfSC)

Characteristics of cell-free DNA (cfDNA) were systematically profiled using key attributes, including fragment length distributions, end motif frequencies, GC content (rounded to two decimal places), nucleosome spacing, and chromosomal origin. Fragments with nucleosome distances greater than 400 bp were aggregated, and those exceeding 450 bp in length were truncated. In total, 1,848 distinct cfDNA features were derived for downstream analyses.

To construct cfDNA scores (cfSC), we employed two distinct feature selection strategies. In the first approach, features were ranked using p-values from the Wilcoxon rank-sum test comparing GDM to CTRL samples in the TRAIN dataset. Features were then selected based on predefined thresholds, including the top 20, 50, 100, 200, 300, 500, 800, 1000, 1500, and all 1,848 features. In the second approach, Gaussian Random Projection was employed to reduce the dimensionality of the feature matrix to 500 components while preserving essential data structure. Subsequently, Hierarchical Density-Based Spatial Clustering of Applications with Noise (HDBSCAN) clustering technique (Campello et al., 2013) was applied to group features by similarity within this reduced space, with the minimum cluster size parameter optimized iteratively. Features identified as noise were excluded from further analysis. For each cluster, the feature exhibiting the greatest absolute mean difference between GDM and control groups was selected as a representative, facilitating the identification of a non-redundant subset strongly associated with the outcome. Following selection of cfDNA structural features, three previously established markers— motif diversity scores (MDS), methylation-associated (MA) value, and fetal fraction—were incorporated into the analysis as described in (Tang et al., 2024). To differentiate among structural scores, each is denoted by the cfSC prefix followed by the number of selected features or the feature selection method employed. For example, cfSC_HDBSCAN_ stands for the cfDNA score in which features are selected via HDBSCAN clustering.

#### Genetics-based score (gbSC)

##### Omics prediction scores and omics selection

A comprehensive atlas of genetic prediction models for approximately 19,000 omics traits was obtained from the OmicsPred database (https://www.omicspred.org/) primarily developed by (Xu et al., 2022). Using this resource, we generated a high-dimensional dataset comprising predicted scores for each trait across all 1,086 individuals in our cohort.

In parallel with the approach employed for cfDNA structural scores, we applied two distinct omics selection strategies. First, we evaluated the association between each omics-derived prediction score and gestational diabetes mellitus (GDM) using the Wilcoxon rank-sum test across our cohort of 1,086 samples.

Omics traits demonstrating statistical significance following Benjamini–Hochberg false discovery rate (FDR) correction were retained for the construction of genetic scores. In a second strategy, we re-assessed the associations between omics traits and GDM within the TRAIN dataset alone. Subsequently, omics traits which are corresponding to the top 20, 50, 100, 200, 300, 500, 800, 1,000, 1,500, 2,000, 2,500, and 3,000 lowest p-values were selected for genetics-based scores development. Each genetics-based score is denoted by the gbSC prefix followed by the number of selected features or the feature selection method employed. For example, gbSC_BH_ stands for the genetics-based score in which omics traits are selected by Bejamin-Hochberg correction.

##### Master score

The master score was constructed by aligning individual component scores using logistic regression, trained on the VALIDATION dataset. Each component score was designed such that higher values indicate an increased likelihood of gestational diabetes mellitus (GDM); accordingly, we expected positive regression coefficients in the final model. We began with an initial logistic regression model including all component scores. To refine the model, we employed a stepwise elimination procedure: at each iteration, a single component score was removed if its regression coefficient was negative or if its *p*-value exceeded 0.05. The performance of the master score is then verified through AUC addition-deletion analysis.

## Result

### Study Design and Workflow

Our study employed a comprehensive multi-modal approach to develop an early GDM prediction model using blood samples collected from 1,086 pregnant women between 9-12 weeks of gestation (Figure 1). The cohort comprised 435 women who were diagnosed with GDM and 651 controls with normal glucose tolerance, with GDM diagnosis confirmed through standard screening protocols conducted between 24-28 weeks of gestation.

**Figure 1:**
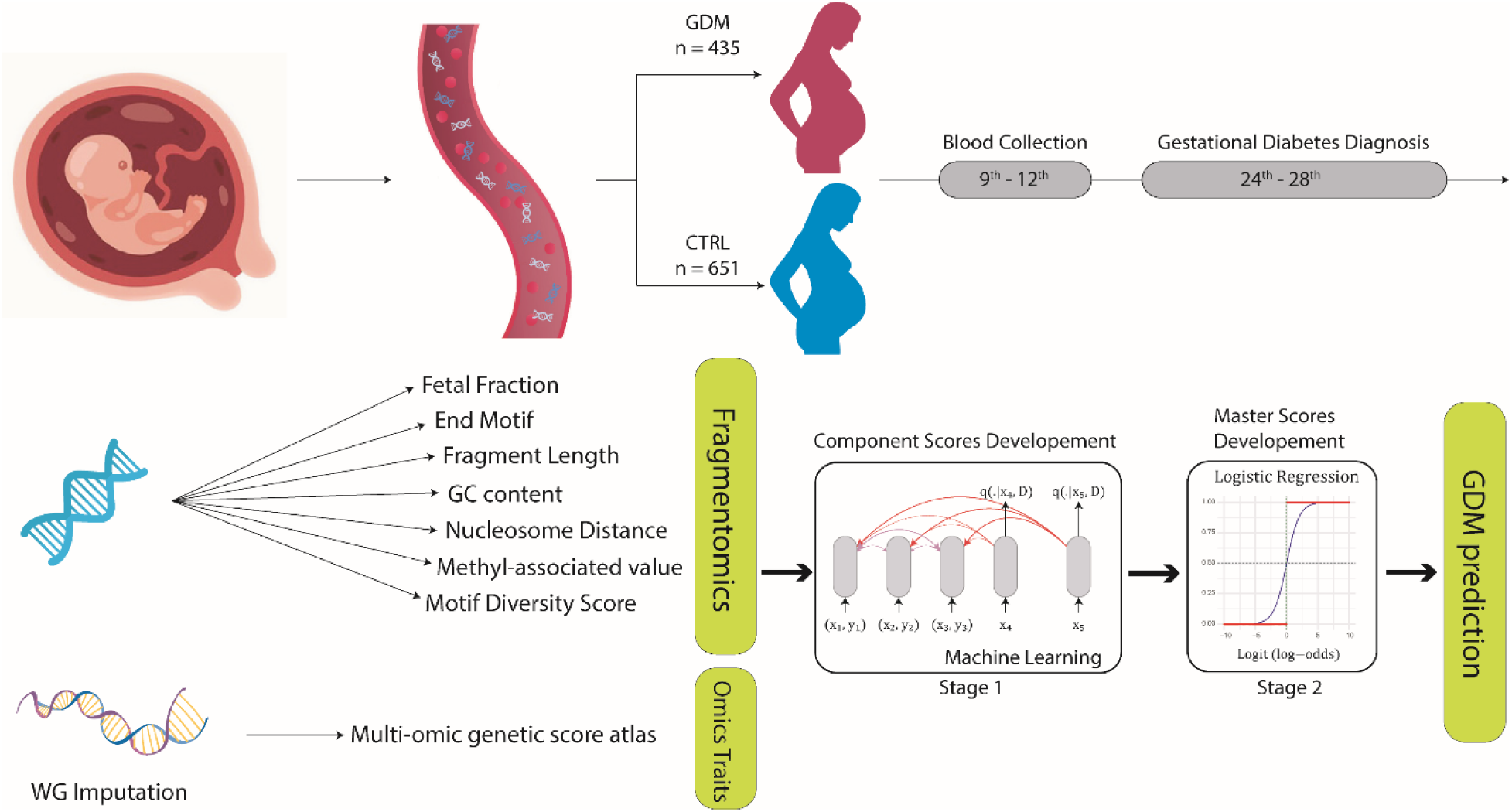
Study design and analytical workflow. Blood samples from 1,086 pregnant women were collected at 9-12 weeks of gestation, with GDM diagnosis confirmed at 24-28 weeks. The cfDNA underwent two parallel processes: cfDNA profiling to extract cfDNA specific features (fetal fraction, end motifs, fragment length, GC content, nucleosome distance, MA, MDS), and whole-genome imputation to derive multi-omic trait prediction scores. A two-stage machine learning framework integrated these complementary data modalities to develop component scores, which were subsequently combined into a unified master score for early GDM prediction.

The analytical workflow incorporated two parallel data generation pathways from circulating cell-free DNA. The first pathway involved cfDNA profiling, extracting cfDNA features including fetal fraction, end motifs, fragment length distributions, GC content, nucleosome distance patterns, along with previously established markers MA and MDS (Tang et al., 2024). These features captured the dynamic, pregnancy-specific characteristics of cfDNA that may reflect ongoing metabolic changes associated with GDM development. The second pathway utilized whole-genome imputation to rebuild individual genetic profiles. These profiles were subsequently used to calculate multi-omics genetic prediction scores for about 19,000 molecular traits, thereby capturing the wider polygenic architecture associated with GDM susceptibility.

These diverse data streams—cfDNA features and genetic information—were integrated using a machine learning framework designed to develop component scores for each data modality before combining them into a unified classification model. This two-stage approach allowed us to harness the complementary strengths of both dynamic cfDNA structural changes and underlying genetic predisposition, ultimately generating a master score optimized for early GDM prediction and clinical risk stratification.

### Omics traits association

Our genome-wide analysis of ∼19,000 omics-derived trait prediction scores across 1,086 samples identified five genes with statistically significant associations with GDM after Benjamini-Hochberg correction for multiple testing (Figure 2). These genes—*HSD11B1*, *NEK7*, *COMMD10*, *KLRC4*, and *OCEL1*—showed the strongest evidence for association with GDM risk, with -log₁₀(p-values) ranging from approximately 4.5 to 6.0. While the Manhattan plot revealed numerous additional signals approaching nominal significance thresholds (P < 0.05 and P < 0.01), the relatively small number of variants surviving multiple testing correction underscores the polygenic nature of GDM, where many genes may contribute modest individual effects. These results suggest the utility of omics-informed predictive models in capturing the complex molecular architecture of GDM and underscore their potential for improving early classification and risk stratification in pregnancy.

**Figure 2:**
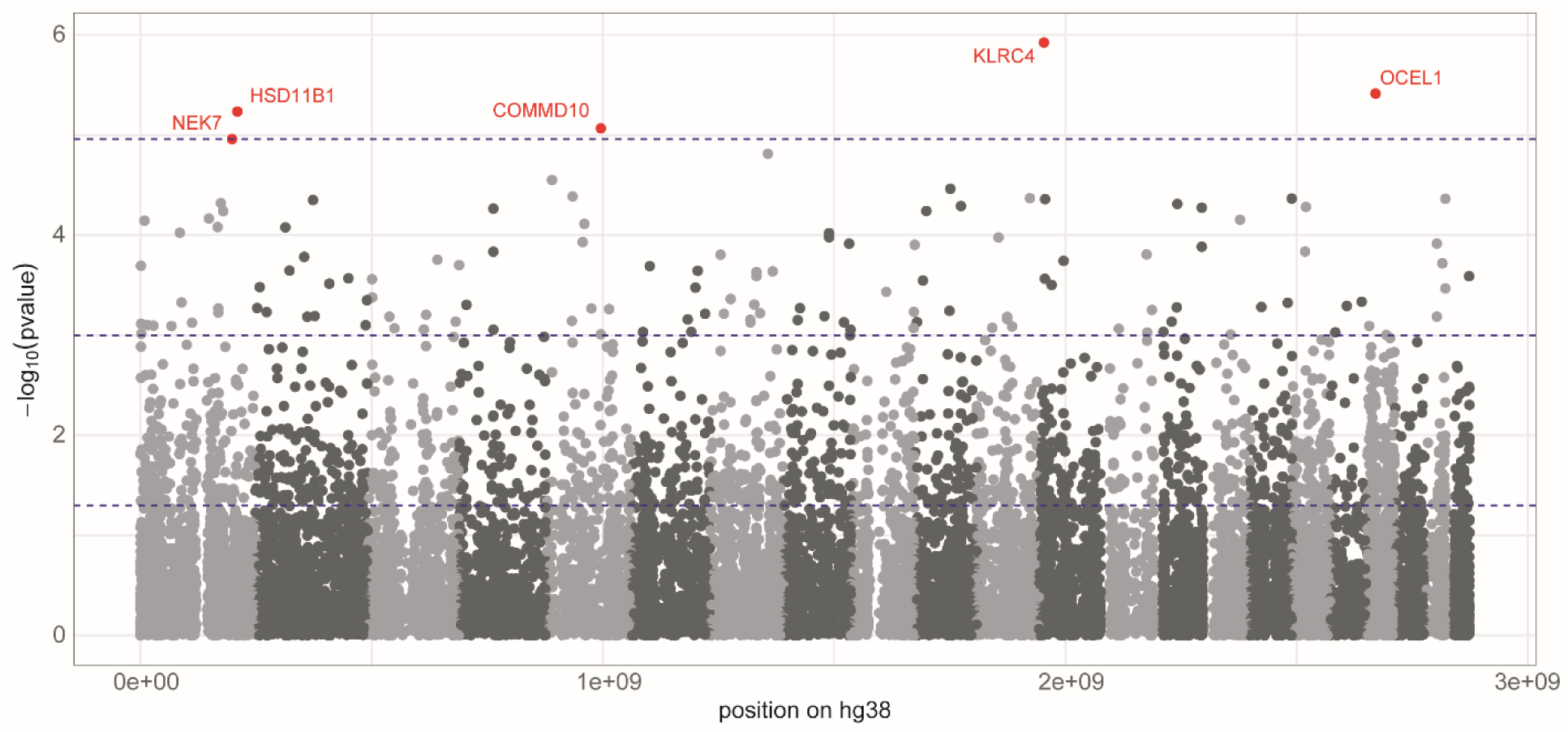
Genome-wide association analysis of omics traits with GDM. Manhattan plot showing the genomic distribution of ∼19,000 omics traits and their association with GDM risk. Each point represents an omics trait positioned by its corresponding gene location or mean SNP location (hg38 assembly). Red points indicate traits showing statistical significance after Benjamini–Hochberg correction (uppermost blue line). Two lower blue lines indicate nominal significance thresholds (P < 0.05 and P < 0.01).

### Component score optimization

The performance of individual component scores demonstrated distinct optimization patterns that varied substantially between cfDNA features and genetics-based predictors (Figure 3 and Table S1). For cfDNA scores (cfSC), we observed a clear performance peak when incorporating 200 features, achieving an AUC of approximately 71.52. Beyond this optimal point, model performance declined progressively, plateauing around 66-68 when including 500 or more features. This performance degradation suggests that adding less discriminative cfDNA structural features introduces noise that outweighs any marginal predictive benefit. Notably, the HDBSCAN-based feature selection approach yielded an AUC of approximately 68.94, which was notably lower than the optimal cfSC_200_ model. This indicates that the unsupervised clustering approach may sacrifice some discriminative power by not directly optimizing GDM classification performance.

**Figure 3:**
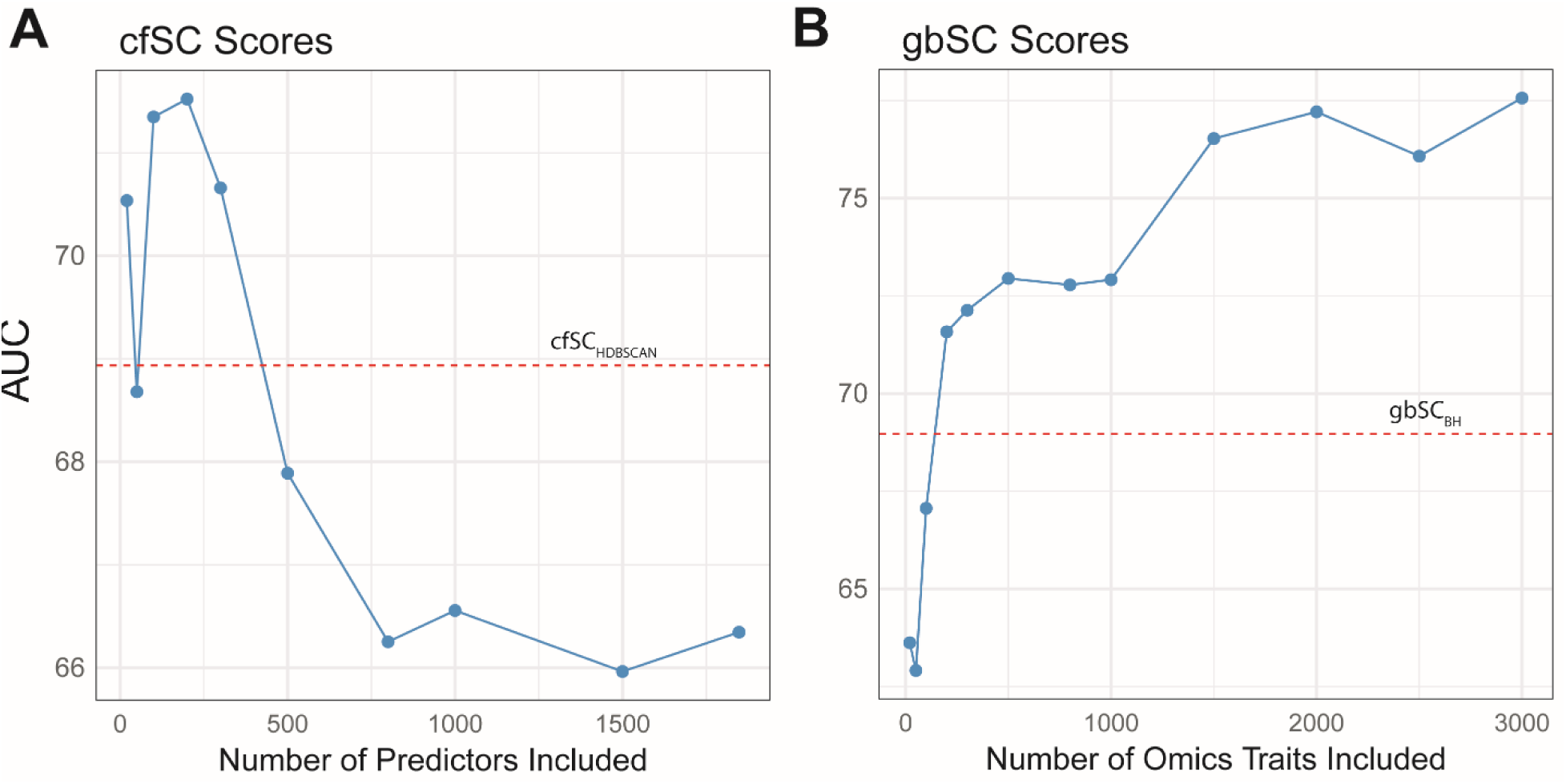
Performance optimization of component scores. AUC values (combined validation and discovery cohorts) plotted against the number of features included in cfSC and gbSC models. Blue curves show performance trends as feature numbers increase. Red dashed lines indicate baseline model performance: cfSC_HDBSCAN_ for cfDNA scores (cfSC) and gbSC_BH_ for genetics-based scores (gbSC). Optimal performance occurs at 200 features for cfSC and about 2,000 features for gbSC.

In contrast, genetics-based scores (gbSC) exhibited markedly different optimization dynamics. Performance improved steadily from an AUC of approximately 63.62 with 20 omics traits to 71.58 with 200 traits. The AUC then remained relatively stable around 72-73 until approximately 800 traits were included, after which performance continued to increase gradually, ultimately reaching peak performance near 77.21 when incorporating 2,000 omics traits.

Notably, the genetics-based score built exclusively from the five statistically significant omics traits (gbSC_BH_) achieved only moderate performance with an AUC of 68.96, substantially lower than models incorporating larger numbers of nominally significant traits. This finding underscores a key insight: while individual omics traits may not reach genome-wide significance thresholds, their collective contribution in a polygenic framework provides substantially greater predictive value than focusing solely on the most statistically significant signals.

### Master score performance

The master score construction process began with an initial logistic regression model incorporating all 24 component scores, but stepwise elimination based on statistical significance and coefficient direction ultimately retained only three components: gbSC_2000_, gbSC_BH_, and cfSC_200_ (Figure 4 and Table S2). This parsimonious final model represents the optimal balance between predictive performance and model complexity, with gbSC_2000_ and cfSC_200_ being the highest-performing representatives from the genetics and structural feature domains, respectively. While individual component scores achieved AUCs ranging from 63 to 77, the master score demonstrated substantially enhanced performance with AUCs of 86.82 (95% CI: 80.98-91.24) in the discovery cohort and 87.19 (95% CI: 81.94-91.72) in the validation cohort. The ROC analysis revealed a sensitivity of 70% (95% CI: 63%-77%) and specificity of 89% (95% CI: 84%-92%) when evaluated across the combined validation and discovery datasets (Figure 4A). Detailed results of the logistic regression are provided in (Table S3).

**Figure 4:**
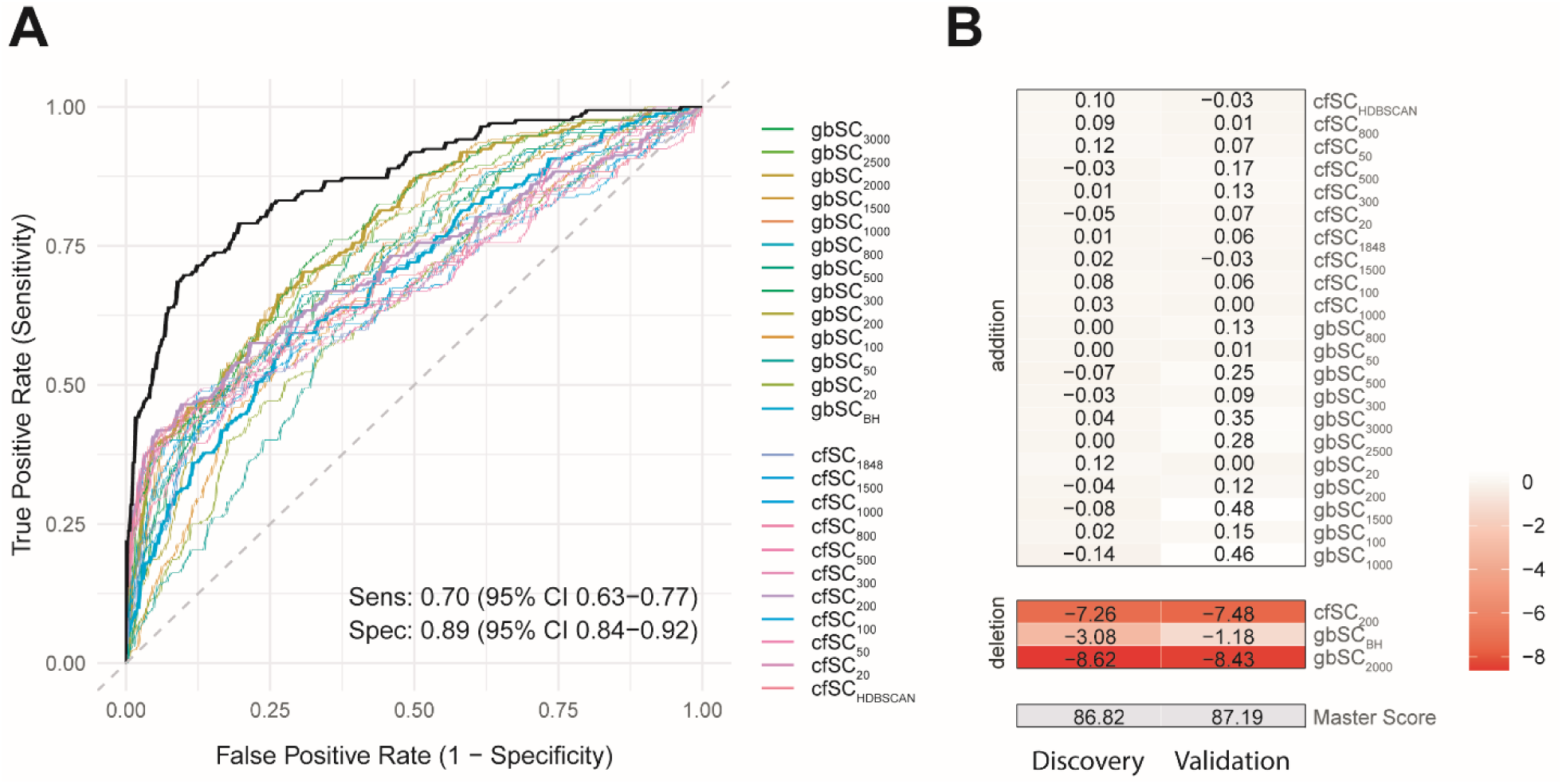
Master score performance and component contribution analysis. (A) AUC values for individual component scores (colored lines) and the integrated master score (black line) in validation and discovery cohorts. Sensitivity and specificity values are shown for the combined validation and discovery datasets. (B) Impact of component score removal (Deletion sub-panel) and addition (Addition sub-panel) on master score AUC. The master score baseline AUC is indicated at the bottom. Removing cfSC_200_ or gbSC_2000_ substantially reduces performance (7-8 percentage points), while adding other components provides minimal improvement.

The addition-deletion analysis provided crucial insights into each component’s contribution to overall performance (Figure 4B). Removal of either gbSC_2000_ or cfSC_200_ from the master score resulted in substantial performance decrements of 7.26 and 8.62 percentage points, respectively, in the validation dataset, with similar magnitudes observed in the discovery cohort. This demonstrates that both genetics-based and cfDNA structural information provide essential, non-redundant contributions to GDM prediction. Interestingly, the removal of gbSC_BH_ showed a more modest impact (1.18 percentage point reduction), suggesting its role may be more complementary than fundamental. Conversely, the addition analysis revealed that incorporating any of the remaining 21 component scores produced minimal improvements in AUC, with most changes being less than 0.5 percentage points. This finding validates the effectiveness of the stepwise elimination procedure and confirms that the three-component master score captures the essential predictive information available in our multi-modal dataset without introducing unnecessary complexity or overfitting.

### Risk Stratification

The master score demonstrated excellent capacity for clinical risk stratification, with clear separation between GDM cases and controls across the full range of score values (Figure 5). The cumulative distribution curves revealed that approximately 88% of controls scored below 0.5, while only 30% of GDM cases fell within this low-risk range. This substantial separation provides a strong foundation for clinical decision-making at early gestational stages.

**Figure 5:**
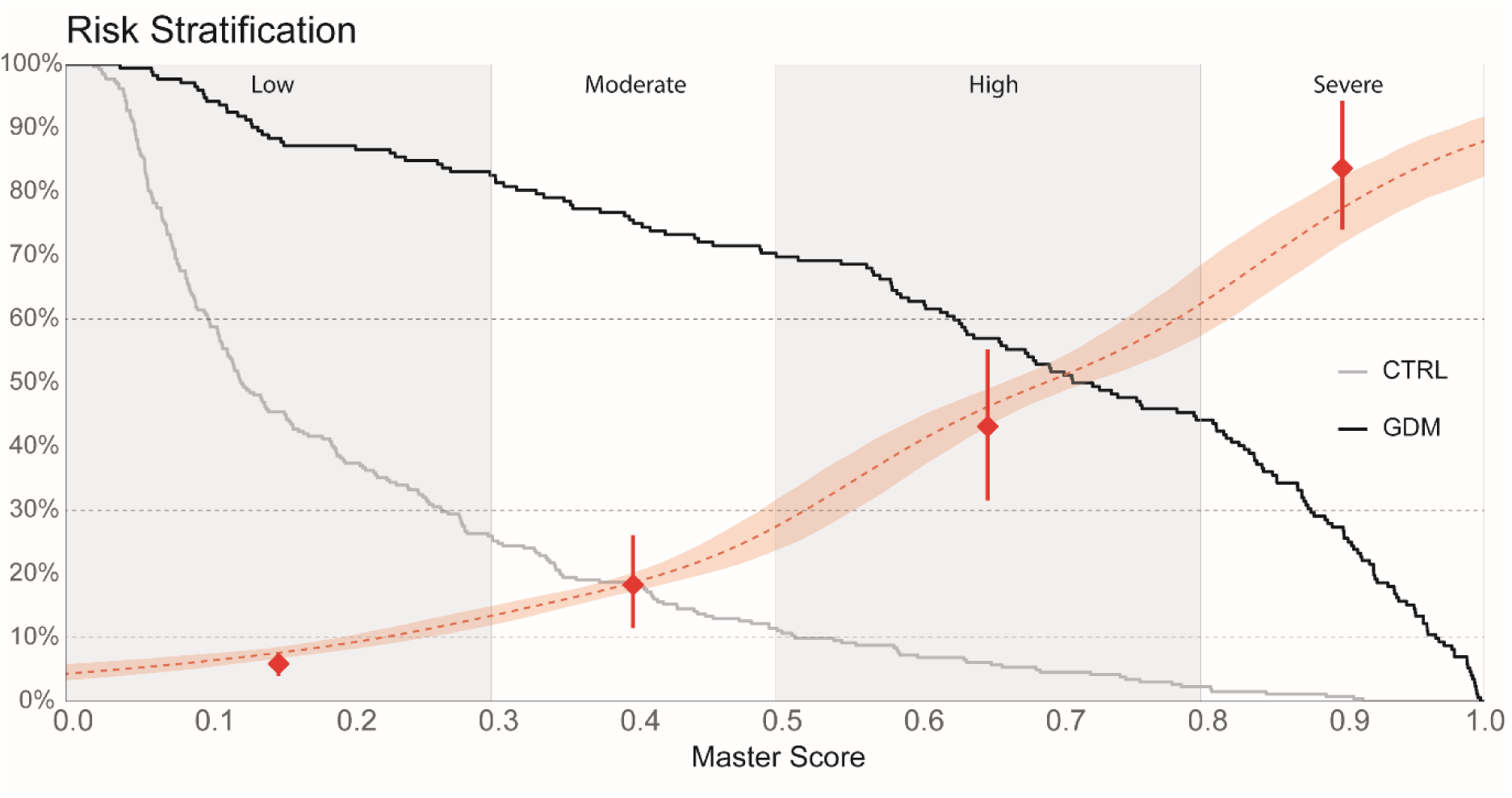
Clinical risk stratification using the master score. Cumulative distribution curves showing the proportion of control (gray) and GDM (black) populations exceeding each master score threshold in combined validation and discovery cohorts. Red dots indicate GDM prevalence within score bins, assuming 21% population prevalence. For example, 75% of GDM cases score above 0.4 compared to 18% of controls, and women scoring above 0.9 have 90% GDM prevalence (95% CI: 79%-100%). The red dashed line shows the relationship between GDM risk and master score.

Using the established Vietnamese population GDM prevalence of 21% as a baseline reference (*Global Prevalence of Gestational Diabetes (GDM) | IDF Atlas*, n.d.), we identified clinically meaningful risk thresholds that could guide prenatal care strategies. Women with master scores of 0.3 or higher demonstrated GDM prevalence rates at least similar to the population baseline. At a cutoff of 0.4, approximately 75% of GDM cases would be identified, while maintaining a relatively low false positive rate of 18% among controls.

The risk stratification analysis revealed four distinct risk categories based on master score ranges. Women scoring between 0.0-0.3 represented a low-risk group with GDM prevalence is below 10%. The moderate-risk category (scores 0.3-0.5) encompassed women with GDM prevalence ranging from 10% to 30%, remaining at similar level of population baseline risk.

Women in the high-risk category (scores 0.5-0.8) showed GDM prevalence of approximately 45%, nearly double the population baseline. Most strikingly, women in the severe-risk category (scores 0.8-1.0) exhibited GDM prevalence exceeding 70%, reaching 90% (95% CI: 78%-100%) for those scoring above 0.9.

## Discussion

The multi-modal machine learning framework developed in this study demonstrates how integrating complementary data types can substantially enhance early GDM prediction compared to single-biomarker approaches. Our findings reveal that genetics-based scores (gbSC) and cfDNA features (cfSC) capture fundamentally different aspects of GDM pathogenesis, with each contributing essential, non-redundant information to the final prediction model.

The genetics-based (gbSC) scores reflect the stable, time-invariant genetic predisposition of pregnant women to GDM development. These scores derive entirely from germline genetic variation and remain constant throughout pregnancy, representing the underlying constitutional risk that each woman carries. In contrast, cfSC scores capture dynamic, pregnancy-specific changes in cfDNA fragmentation patterns that may reflect ongoing metabolic perturbations, placental dysfunction, or inflammatory processes associated with early GDM pathogenesis. The complementary nature of these two data modalities is clearly demonstrated by our addition-deletion analysis, which showed that removing either gbSC_2000_ or cfSC_200_ resulted in substantial performance decrements of 6-8 percentage points.

Importantly, our logistic regression suggests that a substantial proportion of women who will develop GDM exhibit normal cfDNA structural profiles at 12 weeks of gestation but are nevertheless identified through their genetic risk profiles captured by gbSC_2000_. This finding has significant clinical implications, as it indicates that cfDNA structural features alone—while valuable—may be insufficient to capture the full complexity of GDM risk. Future research should explore integrating additional data modalities, including clinical parameters such as maternal age, BMI, and family history (Lyu et al., 2025; Zhu et al., 2025), copy number variation profiles from cfDNA (Y. Wang et al., 2023), and transcription start site accessibility patterns (Tang et al., 2024) to further enhance prediction accuracy.

Several technical limitations warrant consideration. Our imputation approach was restricted to biallelic variants, potentially limiting the completeness of genetic variant representation, particularly for structural variants or rare mutations that might contribute to GDM risk. Additionally, the omics prediction atlas was constructed using the hg19 reference genome while our imputation utilized hg38, creating occasional mismatches that resulted in missing SNPs for some omics trait predictions. Although QUILT2 showed robustness to down sampling (Figure S1), the absence of maternal whole-genome sequencing data prevented direct validation of QUILT2 imputation accuracy within our Vietnamese cohort. Despite these constraints, our classifier demonstrated consistent performance across independent discovery datasets, indicating reasonable generalizability within the target population.

Literature validation of our five genome-wide significant omics traits provides compelling biological support for their roles in GDM pathogenesis. *NEK7*, a critical regulator of *NLRP3* inflammasome activation, shows upregulated expression in GDM pregnancies and correlates with glycemic control markers and systemic inflammation (Kumar et al., 2025). Its expression is further elevated by high glucose exposure, directly linking NEK7 to the inflammatory cascade characteristic of GDM. *HSD11B1*, encoding 11β-hydroxysteroid dehydrogenase type 1, exhibits dysregulated placental expression in GDM, affecting local cortisol metabolism and contributing to the insulin resistance central to disease pathogenesis (Konstantakou et al., 2017; Ondřejíková et al., 2021). COMMD10’s involvement in membrane trafficking and cellular signaling includes modulation of basal insulin secretion, with variants linked to metabolic dysfunction and diabetic vascular complications (L. Zhang et al., 2023). *KLRC4*, encoding a natural killer cell receptor, harbors deleterious variants associated with type 2 diabetes in large-scale genetic studies, suggesting immune-mediated metabolic regulation pathways. *OCEL1* has been consistently identified in genomic screens as a type 2 diabetes susceptibility candidate, though its specific mechanistic role requires further investigation. The convergence of statistical significance and biological plausibility for these genes strengthens confidence in our omics-based approach and suggests that the broader set of 2,000 omics traits likely captures additional relevant biological pathways, even if individual effects are modest.

Our sensitivity analysis illuminates important principles for polygenic disease prediction that extend beyond GDM. While only five omics traits achieved genome-wide significance after multiple testing correction, models incorporating 2,000 nominally significant traits substantially outperformed those built exclusively from statistically significant features. This finding underscores the value of polygenic methods that aggregate modest effects across many loci. The complex polygenic landscape of GDM likely involves thousands of genes contributing small individual effects through interconnected expression networks. Our omics atlas approach effectively reduces the search space from approximately eight million germline SNPs to ∼19,000 interpretable molecular traits, providing a more tractable and biologically meaningful framework for polygenic prediction. This methodology may prove valuable for other complex diseases with substantial polygenic components, including type 2 diabetes, cardiovascular disease, and cancer.

The risk stratification framework developed here offers a practical approach for clinical decision-making, with four well-defined risk categories spanning low (0-10%) to severe (70-90%) GDM probability. Setting a threshold at 0.4 enables identification of 78% of future GDM cases while maintaining an acceptable false positive rate of 18% among controls. This balance appears suitable for a screening context where the goal is early identification to enable preventive interventions. Women classified as moderate risk (scores 0.3-0.5) might benefit from enhanced nutritional counseling, lifestyle modifications, and more frequent monitoring. Those at high risk (scores 0.5-0.8) could warrant earlier glucose tolerance testing, intensified dietary interventions, and consideration of metformin or other pharmacological approaches. The severe-risk women (scores >0.8) might benefit from immediate endocrinology consultation and aggressive management strategies typically reserved for established diabetes. Importantly, this prediction model was designed for apparently healthy pregnant women at 10-12 weeks of gestation and should be interpreted as a probabilistic risk assessment rather than a diagnostic tool. Women classified as high-risk may benefit from preventive interventions that could potentially modify their trajectory and prevent GDM development by 24-28 weeks, when traditional diagnosis occurs.

Vietnam’s exceptionally high GDM prevalence (21%) may reflect unique genetic susceptibility patterns, environmental factors, or gene-environment interactions specific to Southeast Asian populations. Our population-specific model addresses this need while establishing a framework that could be adapted to other high-risk populations with appropriate validation studies.

Future research directions should include: (1) prospective validation in independent Vietnamese cohorts to confirm clinical utility; (2) investigation of the model’s performance in other Southeast Asian populations to assess broader generalizability; (3) integration of additional data modalities including metabolomics, proteomics, and detailed clinical parameters; (4) development of interventional strategies tailored to different risk categories; and (5) cost-effectiveness analyses to guide implementation decisions.

In conclusion, the framework established in this study demonstrates the potential for precision medicine approaches in maternal-fetal medicine, where early identification of high-risk pregnancies could enable targeted interventions to prevent GDM and its associated complications. As sequencing costs continue to decline and omics technologies become more accessible, such multi-modal prediction models may become increasingly feasible for routine clinical implementation, potentially transforming prenatal care from a reactive to a proactive, personalized approach.

## Acknowledgment

We gratefully acknowledge the guidance and valuable contributions of Robert W. Davies and Zilong Li, co-authors of the QUILT2 algorithm, in the development and execution of imputation procedures.

## Author Contributions

The core writing group (DNV, PMD, HG) designed the study, coordinated the multicenter collaboration, performed the data analysis, and drafted the manuscript. All named authors substantially contributed to the interpretation of results, critical revision of the manuscript, and approved the final version for submission. Collaborating investigators at participating sites were responsible for patient recruitment and data collection.

The lead author had full access to all study data and takes responsibility for the integrity of the data and the accuracy of the analysis.

## Data Availability

The data that support the findings of this study are available from the corresponding author upon reasonable request.

## Ethics Statement

Ethical approval for this study was obtained from the appropriate institutional review board. Written informed consent was secured from all participating subjects prior to inclusion in the study. All procedures were performed in compliance with relevant guidelines and regulations to protect the rights and privacy of human participants.

