## Supplementary Figures and Tables for "Early Prediction of Gestational Diabetes Using Integrated Cell-free DNA Features and Omics-derived Genetic Scores"

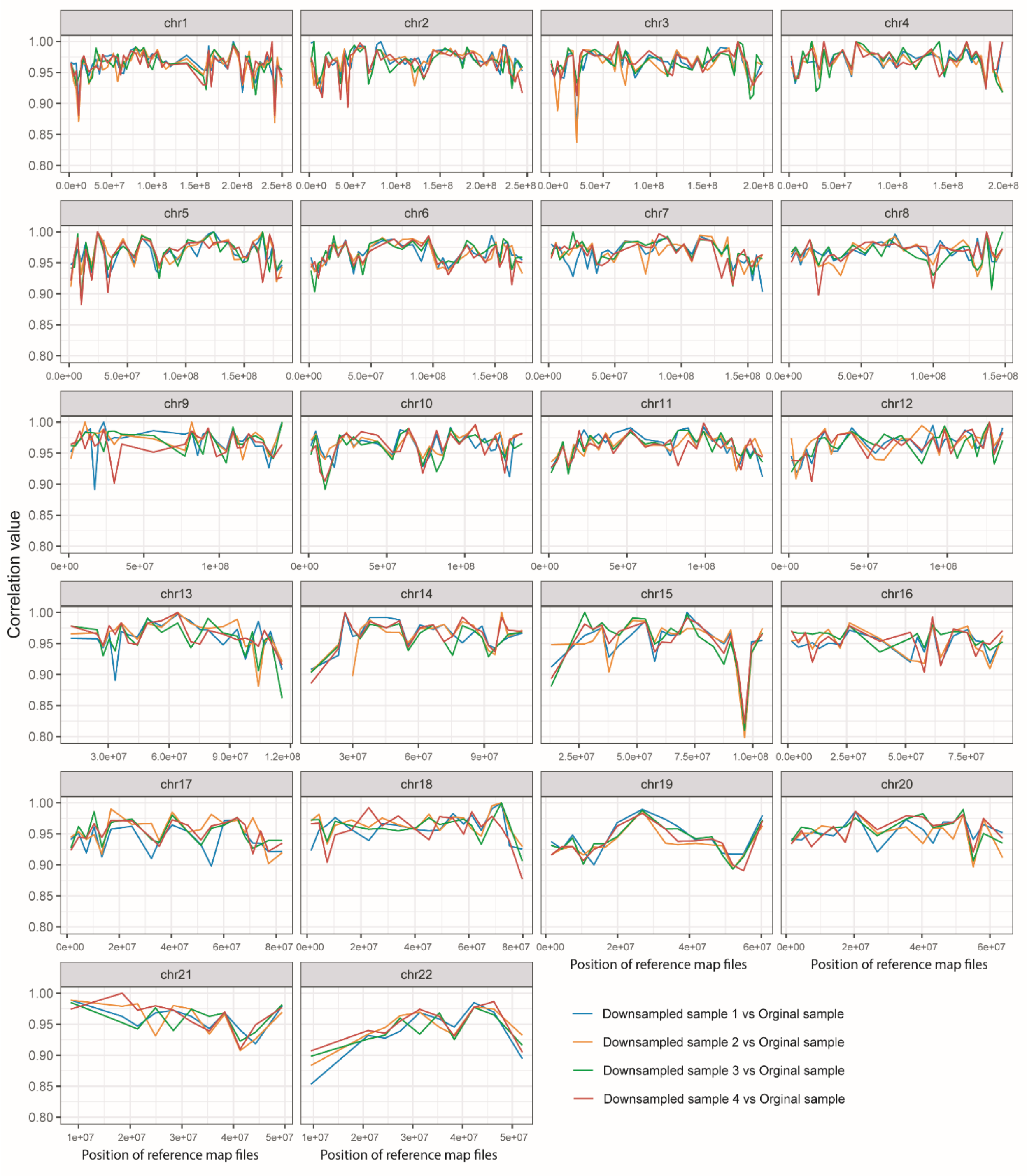


*Figure S1: Imputation robustness was assessed across 616 reference map files (representing 616 regions) for the KHV population. Downsampled datasets were generated from a single original sample (20 million reads) by retaining 30% of the reads. Correlations between the downsampled and original datasets were calculated exclusively for biallelic mutations identified within a pool of 19,000 omics prediction scores. Overall, correlations typically exceeded 0.95.*

### Tables

*Table S1: Performance of component scores assessed by AUC across TRAIN, VALIDATION, and DISCOVERY datasets.*

| Component Scores | TRAIN | VALIDATION+DISCOVERY | VALIDATION | DISCOVERY |
|---|---|---|---|---|
| $cfSC_{HDBSCAN}$ | 94.35 | 68.94 | 72.34 | 65.70 |
| $cfSC_{20}$ | 75.38 | 70.54 | 71.46 | 69.80 |
| $cfSC_{50}$ | 78.82 | 68.68 | 69.32 | 68.09 |
| $cfSC_{100}$ | 86.94 | 71.35 | 74.78 | 67.47 |
| $cfSC_{200}$ | 89.99 | 71.52 | 75.76 | 67.71 |
| $cfSC_{300}$ | 92.55 | 70.66 | 75.41 | 66.18 |
| $cfSC_{500}$ | 95.16 | 67.89 | 71.18 | 64.66 |
| $cfSC_{800}$ | 97.73 | 66.25 | 65.99 | 66.71 |
| $cfSC_{1000}$ | 98.12 | 66.56 | 69.32 | 64.02 |
| $cfSC_{1500}$ | 98.81 | 65.96 | 69.26 | 62.46 |
| $cfSC_{1848}$ | 98.57 | 66.35 | 70.64 | 62.19 |
| $gbSC_{BH}$ | 78.13 | 68.96 | 68.10 | 69.69 |
| $gbSC_{20}$ | 90.22 | 63.62 | 63.93 | 63.38 |
| $gbSC_{50}$ | 95.47 | 62.91 | 62.42 | 63.59 |
| $gbSC_{100}$ | 98.15 | 67.06 | 65.08 | 68.79 |
| $gbSC_{200}$ | 99.86 | 71.58 | 72.76 | 70.39 |
| $gbSC_{300}$ | 99.98 | 72.13 | 72.78 | 71.06 |
| $gbSC_{500}$ | 100.00 | 72.95 | 72.19 | 73.50 |
| $gbSC_{800}$ | 100.00 | 72.78 | 73.10 | 72.65 |
| $gbSC_{1000}$ | 100.00 | 72.92 | 71.92 | 73.91 |
| $gbSC_{1500}$ | 100.00 | 76.53 | 76.85 | 76.60 |
| $gbSC_{2000}$ | 100.00 | 77.21 | 77.87 | 76.64 |
| $gbSC_{2500}$ | 100.00 | 76.08 | 76.29 | 76.09 |
| $gbSC_{3000}$ | 100.00 | 77.57 | 77.79 | 77.39 |

*Table S2: Logistic regression summary.*

| **Predictors** | **Coefficient** | **Standard error** | **z-value** | **P-value** |
|---|---|---|---|---|
| **Intercept** | -5.4129 | 0.7752 | -6.983 | 2.89e-12 |
| **$gbSC_{2000}$** | 3.0631 | 0.5495 | 5.569 | 2.56e-08 |
| **$gbSC_{BH}$** | 3.4779 | 1.2530 | 2.776 | 0.00551 |
| **$cfSC_{200}$** | 6.2750 | 1.1357 | 5.525 | 3.29e-08 |
